# Two-year recall for people with no diabetic retinopathy: a multiethnic population-based retrospective cohort study using real-world data to quantify the effect

**DOI:** 10.1101/2023.06.14.23291369

**Authors:** Abraham Olvera-Barrios, Alicja R Rudnicka, John Anderson, Louis Bolter, Ryan Chambers, Alasdair N Warwick, Roshan Welikala, Jiri Fajtl, Sarah Barman, Paolo Remagnino, Yue Wu, Aaron Lee, Emily Y Chew, Frederick L. Ferris, Aroon D Hingorani, Reecha Sofat, Catherine Egan, Adnan Tufail, Christopher G Owen, the ARIAS Research Group

**Affiliations:** University College London Institute of Ophthalmology, London, UK; Moorfields Eye Hospital NHS Foundation Trust, London, UK; Population Health Research Institute, St. George’s, University of London, London, UK; Homerton Healthcare NHS Foundation Trust, London, UK; Institute of Cardiovascular Science, University College London, London, UK; School of Computer Science and Mathematics, Kingston University, London, UK; Department of Computer Science, University of Durham, Durham, UK; Department of Ophthalmology, University of Washington, Seattle, WA, USA; Roger and Angie Karalis Johnson Retina Center, Seattle, WA, USA; Division of Epidemiology and Clinical Applications, National Eye Institute, National Institutes of Health, Bethesda, Maryland, USA; Ophthalmic research consultants, Charlotte, North Carolina, USA; Department of Pharmacology and Therapeutics, University of Liverpool, UK

**Author notes:** **Correspondence:** John Anderson (email address), Christopher G Owen (email address). **Synopsis/Precis** Introduction of biennial diabetic eye screening among people living with diabetes with no diabetic retinopathy on 2 annual screening appointments could accentuate sociodemographic differences in diabetes related sight loss, especially among younger and non-white populations.

## Abstract

**Background/Aims:** The English Diabetic Eye Screening Programme (DESP) offers people living with diabetes (PLD) annual screening. Less frequent screening has been advocated among PLD without diabetic retinopathy (DR), but evidence for each ethnic group is limited. We examined the potential effect of biennial vs annual screening on the detection of sight-threatening diabetic retinopathy (STDR) and proliferative diabetic retinopathy (PDR) among PLD without DR from a large urban-multi-ethnic English DESP.

**Methods:** PLD in North-East London DESP (Jan-2012 to Dec-2021) with no DR on two prior consecutive screening visits with up to eight years of follow-up were examined. Annual STDR and PDR incidence rates, overall, and by ethnicity were quantified. Delays in identification of STDR and PDR events had 2-year screening intervals been used were determined.

**Findings:** Among 82,782 PLD (37% white, 36% South Asian, and 16% black people), there were 1,788 incident STDR cases over mean 4.3 (SD 2.4) years (STDR rate 0.51, 95%CI 0.47-0.55 per 100-person-years). STDR incidence rates per 100-person-years by ethnicity were 0.55 (95% CI 0.48-0.62) for South Asian, 0.34 (0.29-0.40) for white, and 0.77 (0.65-0.90) for black people. Biennial screening would have delayed diagnosis by 1-year for 56.3% (1,007/1,788) with STDR and 43.6% (45/103) with PDR. Standardised cumulative rates of delayed STDR per-100,000 for each ethnic group were 1904 (95%CI 1683-2154) for black, 1276 (1153-1412), and 844 (745-955) for white people.

**Interpretation:** Biennial screening would have delayed detection of some STDR and PDR by one-year especially among those of black ethnic origin, leading to healthcare inequalities.

**Key messages:** *What is already known on this topic?:* The UK National Screening Committee currently recommends annual eye screening for diabetic retinopathy among people living with diabetes at high risk of sight loss, but biennial screening among those at low risk of sight loss. Ethnic differences in diabetes and the development of sight-threatening diabetes complications have been reported. The effect of biennial vs annual diabetic eye screening among different ethnic groups at low risk of complications has not been quantified in large multi-ethnic diabetic eye screening programmes in the UK.

*What this study adds?:* We provide incidence rates for the development of new sight-threatening diabetic retinopathy and proliferative diabetic retinopathy in a low-risk group, overall and by different ethnic and age groups, in this diverse sociodemographic population without previous diabetic retinopathy. Implementation of biennial screening in this population would have delayed referral to hospital eye services by a year in near half of those with sight-threatening diabetes (56%) and proliferative retinopathy (44%), but higher absolute rates of delay were observed among the youngest and oldest compared with middle aged and pre-retirement age groups, and those of black ethnic origin compared with other ethnic groups. Higher hazards of STDR were observed in younger people. While the absolute number delayed is small relative to the size of the overall cohort, age and ethnic inequalities in delayed identification of complications were apparent.

*How this study might affect research, practice or policy?:* National implementation of a 2-year diabetic eye screening interval for people with low-risk diabetic retinopathy grades does not affect all population sub-groups equally with respect to delays in the detection and referral of the most serious eye disease. Younger people and people of black and Asian ethnicities are affected more than other groups with potential effects on vision and treatment outcomes.

## Introduction

Diabetic retinopathy is a major microvascular complication of diabetes which can result in sight loss, presenting a major global challenge.^1^ However, early detection and treatment can prevent or delay sight loss. The NHS Diabetic Eye Screening Programme (DESP) was introduced in 2003 to identify those with diabetic retinopathy so early treatment can be used.^2^ The English NHS DESP currently performs 2.3 million eye screening appointments each year, generating approximately 13 million retinal images, and the number of appointments and images has increased over time.^3^ Retinal images from the DESP are assessed by up to 3 trained human graders for the presence and severity of diabetic retinopathy (DR), and those with potentially sight-threatening diabetic retinopathy (STDR) are referred to Hospital Eye Services for further assessment and potential treatment. This represents a major challenge to healthcare providers, given increasing patient numbers and finite resources within a publicly funded healthcare system.

Evidence has suggested that biennial rather than annual screening among those at low risk would be safe and cost-effective, potentially reducing the number of appointments and workload.^4-6^ However, not all evidence has been as supportive, concluding that there is insufficient evidence to recommend screening beyond one year.^7^ The UK National Screening Committee (NSC) recommended change in 2016 to biennial screening for those at low risk of sight loss.^8^ The rationale for change was predominantly based on an audit commissioned by the NSC of nearly 350,000 patients from 7 geographically dispersed UK DESPs. This showed progression to STDR (and more serious proliferative diabetic retinopathy [PDR]) among those without DR at two successive screening episodes at least 12 months apart was low (with approximately 0.7% developing referable DR over 2 years).^9^ While this number was considered low, a number of limitations were raised, including the use of retrospective audit data (as opposed to use of preferred randomised controlled trial data), and whether extending follow-up to 2 years could adversely impact attendance once introduced, especially among some sociodemographic groups.^9^ Moreover, while use of the geographically dispersed UK DESP centres would incorporate different age, sex and ethnicity profiles, effects of biennial screening by ethnicity and different age groups were not explicitly quantified. This is highly relevant for ethnicity, given ethnic differences in both diabetes and complications of diabetes, particularly in a UK setting, where those of South Asian ethnicity are at higher risk of diabetes, severe diabetic retinopathy and associated sight-loss, compared with white.^10-12^ While biennial screening among those at low risk of sight loss has been approved, uptake thus far been limited (despite the potential resource and cost savings). Hence, it remains unclear whether this extended screening frequency could lead to inequalities in healthcare.

Using one of the largest, most ethnically diverse DESP in North-East London (NELDESP), we examined progression to STDR and PDR among those without DR on two consecutive annual screens to determine incidence rates by sociodemographic groups, and the potential for delay in the detection of STDR and more serious PDR if biennial screening was introduced, rather than current annual screening interval.

## Methods

The study population comprised people living with diabetes (PLD) registered in the NELDESP, who were offered screening appointments from 03-January-2012 to 31-December-2021. The study was approved by the NHS Health Research Authority and Health and Care Research Wales (IRAS No. 265637). The study was carried out in accordance with the Declaration of Helsinki.

### Setting

North-East London is an ethnically diverse region with higher than national average levels of deprivation and mortality.^13^ The NELDESP is provided by the Homerton Healthcare NHS Foundation Trust, and serves people with diabetes living in inner-city areas with multi-ethnic populations. The NELDESP is run according to English NHS DESP standards.^14^ People with diabetes aged ≥12 years are identified through the electronic ‘General Practice to Diabetic Retinopathy Screening’ patient-identification system. This notifies DESPs about all people with diabetes in their catchment area. All new eligible people are invited for screening within 3 months of notification and the list of PLD eligible for screening by NELDESP is actively maintained.^15^

### Screening visit

A screening visit entails history taking by specialist staff, visual acuity assessment, and capture under pupil dilation of two 45° digital retinal images, centred on the fovea and optic nerve for each eye, respectively. Up to 3 qualified graders assess the images for presence and severity of diabetic retinopathy following a multilevel internally and externally quality-assured process.^14^ The UK National Screening Committee (NSC) classification system for diabetic retinopathy grades in order of increasing severity follows: no retinopathy (R0), mild non-proliferative diabetic retinopathy (R1), severe non-proliferative diabetic retinopathy (R2), diabetic maculopathy (M1), and PDR (R3).^16^ STDR comprises retinopathy grades R2, M1, and R3 and referred according to NSC timescales to hospital eye services for assessment and potential treatment; PDR is urgently referred. Images which were not able to be graded (U) were excluded from the analysis.

### Data extraction

We identified people registered in the NELDESP during the study period, calculated post code-derived index of multiple deprivation (IMD) rank scores for each episode and carried out an anonymised data extraction for all appointments using structured query language searches. An anonymised database was created and stored within the Homerton Trust’s network for analysis. The cohort went through a staged exclusion process illustrated in Supplementary Figure 1, to identify a study cohort of PLD with 2 consecutive annual screening episodes grades of no DR (i.e., R0M0) in both eyes.

### Variables

Routinely collected data from the NELDESP included age at first appointment (categorised as <45, 45 to <55, 55 to <65, and 65 years and older), sex, self-defined ethnicity (coded according to Office for National Statistics [ONS] standards as: white, black, South Asian, Chinese, any other Asian, mixed, other, and unknown categories for the purpose of these analyses),^17^ type of diabetes (Type 2, Type 1, other, and unknown), self-defined duration of diabetes or from date of diagnosis as registered on the screening database, baseline retinopathy severity (to identify those with no diabetic retinopathy [R0M0] in either eye on 2 consecutive screening visits),^18^ and IMD. The IMD combines and weights indicators of deprivation and is the nationally recognised measure of relative deprivation in England.^19^ IMD scores were split into quintiles (where 1^st^ and 5^th^ are the most and least deprived, respectively) following data of the 2019 English indices of deprivation.^19^

### Statistical analysis

We calculated annual incidence rates of STDR (defined as presence of any R2, R3, or M1) in either eye,^16,18^ among those with 2 consecutive annual screening visits without DR (R0M0). Rates were reported by age group, sex, and ethnic group. Note, the median follow-up period between appointments was 1.0 (0.9-1.1) years, providing annual rates. Mutually adjusted hazard ratios for the development of STDR were calculated using Cox regression by age group, sex, ethnic, IMD groups and by duration of diabetes. The proportionality assumption was assessed by graphical inspection of Schoenfeld residuals. To examine the impact of biennial screening intervals, PLD who were R0M0 on two consecutive annual screens were assigned to a virtual biennial screening schedule. Fourteen-month time breaks were used to mirror the annual cycle uptake observed in this cohort. The number of STDR and PDR occurring between biennial screening intervals was quantified. People who developed DR (grades ≥R1M0) were right censored at the screening visit when DR was detected. All analyses were undertaken with R (version 4.2.2).^20^ The Survival^21^ package was used for survival analyses.

## Results

A total of 82,782 PLD from an identified cohort of 200,304 PLD in the NELDESP remained with all relevant demographic data, eyes which could be adequately assessed using fundus photography, and who had no prior diabetic retinopathy on 2 consecutive screening occasions (Supplemental Figure 1). Table 1 summarises baseline characteristics of the cohort where mean age at baseline was 56.7 (SD 14.4) years and 52% (n/N=42,846/82,782) were male.

**Table 1:** Baseline characteristics of the cohort among those with no DR on two consecutive annual screening visits.

| **Characteristic** | **N = 82,782 (%)** |
| --- | --- |
| **Follow-up (SD) years** | 4.3 (2.4) |
| **Age at baseline (SD) years** | 56.7 (14.4) |
| **Age categories** |  |
| <45yr | 16,488 (20%) |
| 45 to <55yr | 20,207 (24%) |
| 55 to <65yr | 20,762 (25%) |
| 65yr and over | 25,325 (31%) |
| **Sex** |  |
| Female | 39,936 (48%) |
| Male | 42,846 (52%) |
| **Type of diabetes** |  |
| Type 2 | 78,992 (95%) |
| Type 1 | 2,125 (2.6%) |
| Other | 137 (0.2%) |
| Missing | 1,528 (1.8%) |
| **Ethnicity** |  |
| White | 30,350 (37%) |
| South Asian | 29,703 (36%) |
| Black | 13,391 (16%) |
| Any other Asian | 4,786 (5.8%) |
| Other | 2,319 (2.8%) |
| Mixed | 1,006 (1.2%) |
| Chinese | 577 (0.7%) |
| Unknown | 650 (0.8%) |
| **Duration of diabetes** | 4.0 (5.3) |
| **Index of Multiple Deprivation** |  |
| 1 | 8,855 (11%) |
| 2 | 26,255 (32%) |
| 3 | 23,956 (29%) |
| 4 | 15,510 (19%) |
| 5 | 8,192 (9.9%) |
| Missing | 14 (<0.1%) |

**Figure 1:**
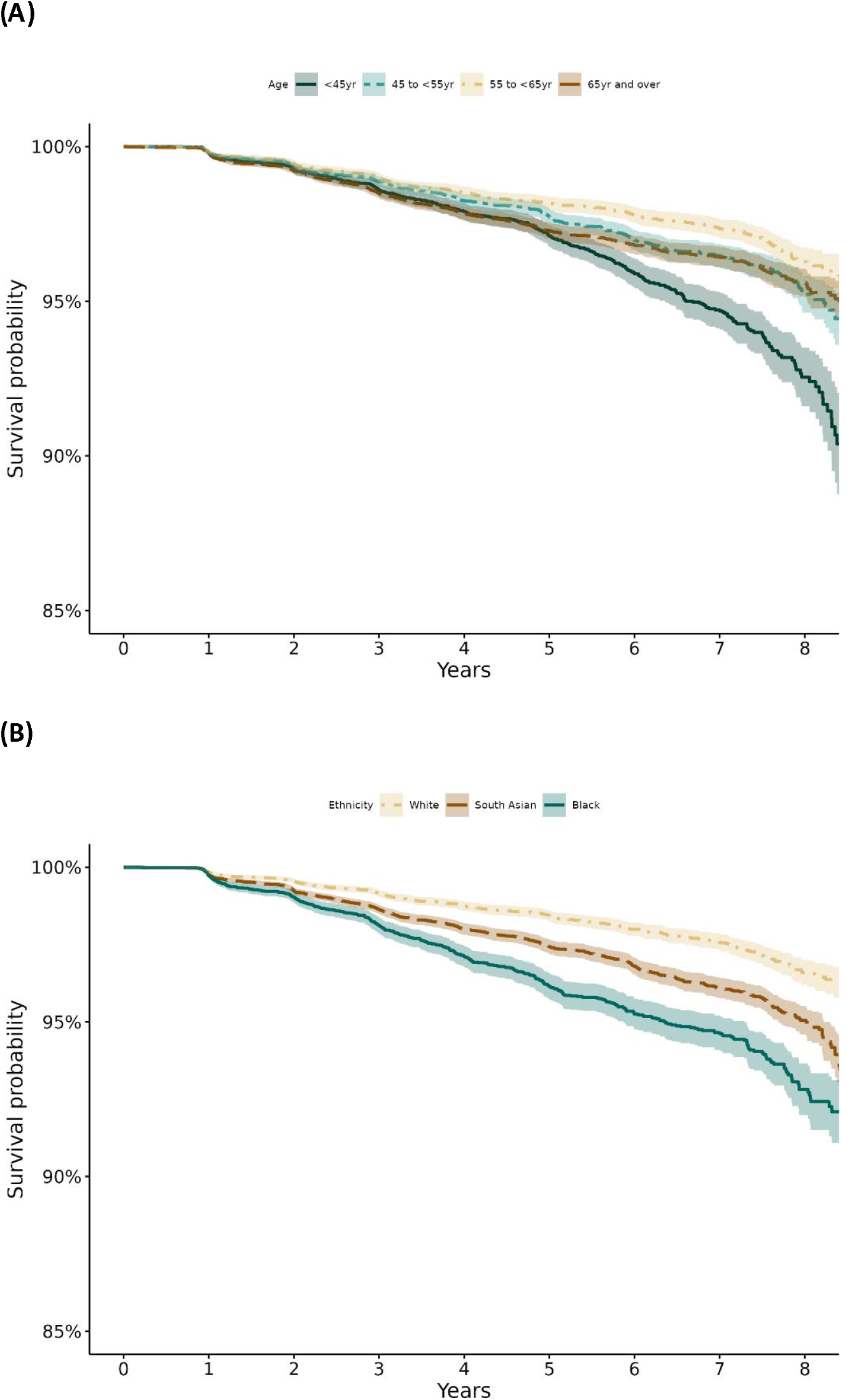
Kaplan Meir plots showing probability of STDR survival over time by (A) age and (B) ethnic group.

Cumulative incidence rates of STDR over the follow-up period are shown in Supplemental Table 1, by age, sex, ethnicity, type of diabetes and IMD group. Progression to STDR with advancing yearly intervals showed a graded increase in cumulative rates over time, which was more pronounced in the youngest and oldest age groups when compared with lower rates in middle age and pre-retirement age groups. Males had consistently lower STDR rates compared with females, and those with Type 1 diabetes consistently higher rates compared with Type 2 diabetes, reflective of diabetes duration. There was no clear pattern across levels of deprivation (IMD). The most striking differences in STDR rates over time were with ethnicity, where PLD of black ethnicity had the highest STDR rates, with South Asian and any other Asian having higher rates compared with white. Those categorised as ‘mixed’ or ‘other’ ethnicity also showed higher rates over the study period. These sociodemographic differences in STDR rates were confirmed by hazard ratios (Table 2) showing markedly higher risk of STDR among black people (121% higher, 95%CI 93-153%) and modestly higher risk among South Asian individuals (54% higher, 95%CI 35-74%) compared with white people. The decreased risk of STDR with increasing age (with the lowest risk among those of pre-retirement age compared with the youngest age group) is also apparent. Sex was not associated. Figure 1 shows Kaplan-Meier plots of STDR survival which shows that the probability of remaining STDR-free over the 8-year study period is lowest among the youngest age group and highest among the pre-retirement age group (Figure 1A), lowest among black individuals, intermediate among South Asian people, and highest among white people (Figure 1B).

**Table 2:** Mutually adjusted hazard ratios of STDR by age groups, sex, ethnic group and IMD in those with two consecutive screening appointments with no retinopathy (R0M0)

| Characteristic | HR (95% CI) | p-value |
| --- | --- | --- |
| **Age (per 5-year increase)** | 0.96 (0.94, 0.98) | **1.5e-4** |
| **Age categories** |  |  |
| <45yr | 1.00 |  |
| 45 to <55yr | 0.70 (0.61, 0.80) | **2.6e-07** |
| 55 to <65yr | 0.54 (0.47, 0.63) | **2.8e-16** |
| 65yr and over | 0.70 (0.61, 0.81) | **1.2e-06** |
| **Sex** |  |  |
| Female | 1.00 |  |
| Male | 0.96 (0.88, 1.06) | 0.449 |
| **Ethnicity** |  |  |
| White | 1.00 |  |
| South Asian | 1.54 (1.35, 1.74) | **2.4e-11** |
| Black | 2.21 (1.93, 2.53) | **8.5e-31** |
| Any other Asian | 1.39 (1.12, 1.72) | **0.003** |
| Other | 1.86 (1.42, 2.44) | **6.1e-06** |
| Mixed | 2.07 (1.39, 3.07) | **3.3e-04** |
| Chinese | 0.63 (0.26, 1.52) | 0.306 |
| Unknown | 1.91 (0.85, 4.28) | 0.118 |
| **Duration of diabetes (per 5-year increase)** | 1.26 (1.21, 1.30) | **1.1e-38** |
| **Type of diabetes** |  |  |
| Type 2 | 1.00 |  |
| Type 1 | 1.41 (1.08, 1.83) | **0.011** |
| Other | 1.83 (0.68, 4.88) | 0.229 |
| Missing | 1.35 (0.91, 2.01) | 0.134 |
| **Deprivation (IMD quintiles)** |  |  |
| 1 | 1.00 |  |
| 2 | 1.16 (0.98, 1.37) | 0.081 |
| 3 | 1.06 (0.90, 1.26) | 0.487 |
| 4 | 1.08 (0.90, 1.30) | 0.393 |
| 5 | 1.12 (0.91, 1.39) | 0.288 |

We examined the potential impact of a biennial screening pathway. The numbers that developed STDR and PDR in the intervening years overall and by ethnic group are shown in Figure 2. Among the 82,782 PLD, STDR was present in 1,788 and PDR in 103 over the study period. However, if the cohort had undergone biennial screening, STDR and PDR would have been present in 56.3% (1,007/1,788) and 43.6% (45/103) at the one-year interval, respectively (Figure 2). Hence, there would have been a one-year delay in the diagnosis of these cases with biennial screening. The near 50% with a one-year delay in STDR and PDR diagnosis remained consistent over the study period (Figure 2). The delayed STDR cases by ethnic group were 256/30,350 for white, 379/29,730 for South Asian, and 256/13,391 for black individuals (Table 3); equivalent to 844, 1276, and 1904 per 100,000 screened biennially, for each ethnic group, respectively (Table 3). For PDR, numbers were much lower, but rates were still higher among black people (90 per 100,000), compared with white (46 per 100,000) and South Asian individuals (44 per 100,000). By age group, delayed STDR events per 100,000 persons were highest (1504 events, 95%CI 1327-1705) among those age <45 years, 1178 (95%CI 1036-1339) for those aged 45 to <55 years, lowest (987 events, 95%CI 859-1134) among those age 55 to <65 years, and 1248 (95%CI 1116-1394) in the oldest age group aged 65 years and over (Table 3). For PDR, there were fewer PDR events among the youngest age groups (36 per 100,000), but markedly more among the oldest age group (95 per 100,000).

**Table 3:**
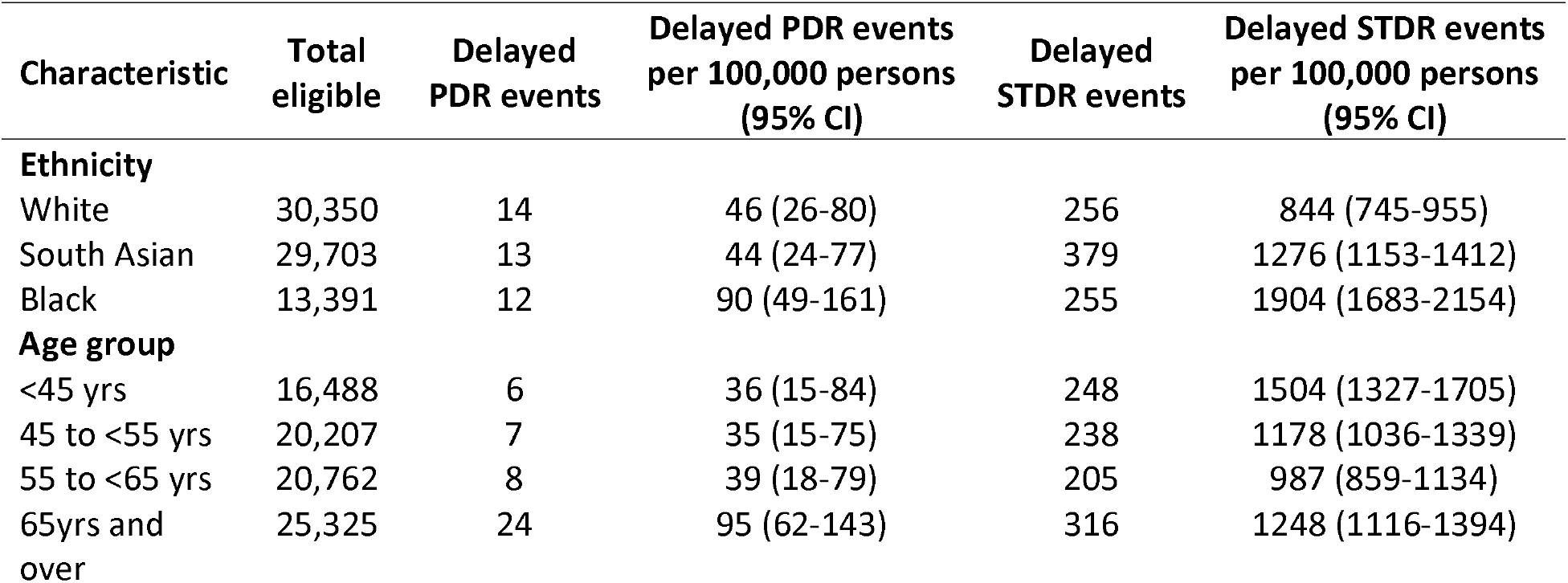
Numbers with delayed PDR and STDR events associated with biennial screening by ethnic and age group.

**Figure 2:**
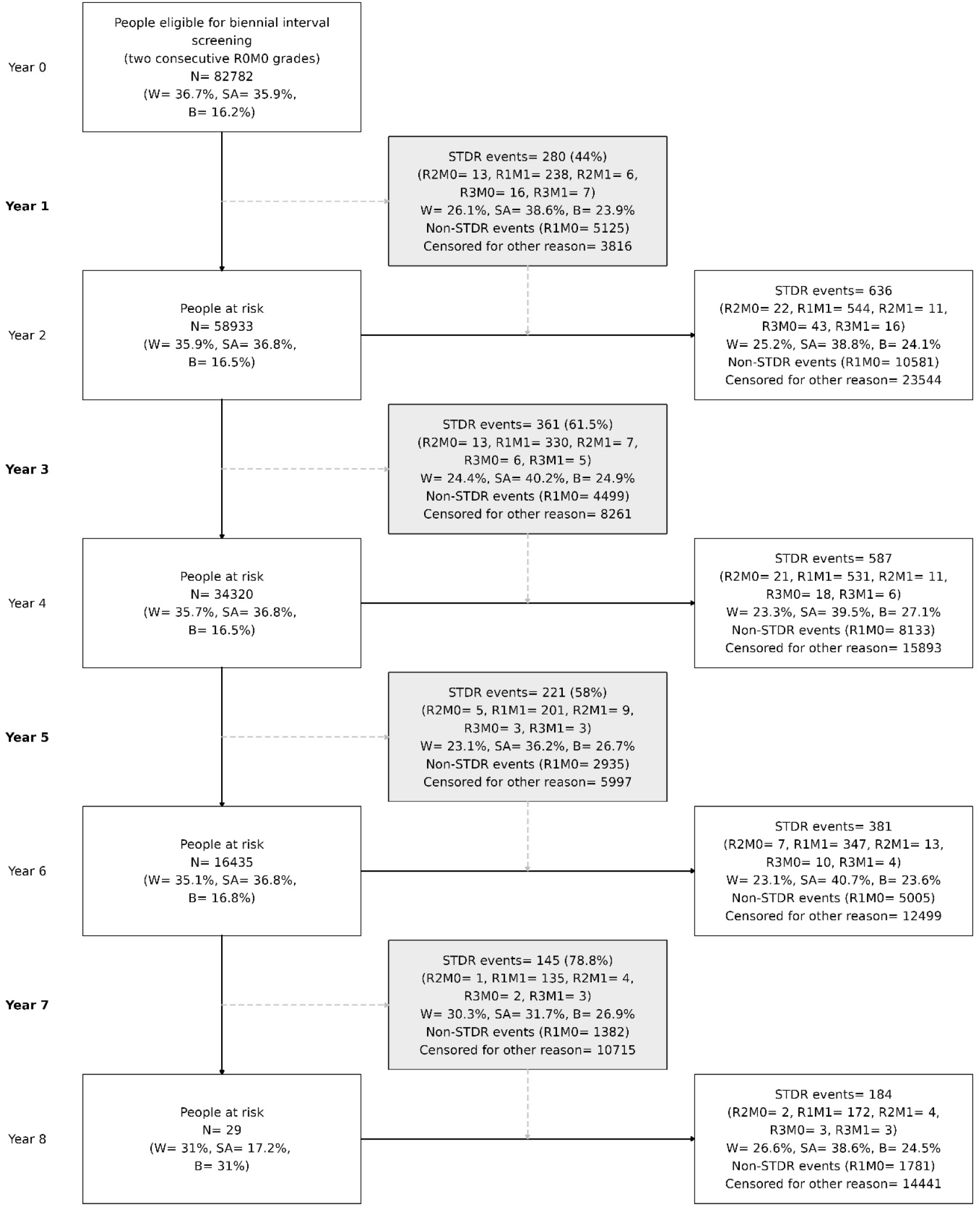
Model of biennial screening in the NELDESP cohort. Grey boxes in column 2 show the number of STDR and PDR events that would have been diagnosed and referred in routine annual screening during the biennial interval but diagnosed at least one year later in two-yearly interval screening. A breakdown by 3 major ethnic groups is presented in each box. Percentages in grey boxes relative to the total STDR and PDR (R3) events from a 2-year interval. W: white, SA: South Asian, B: black.

## Discussion

Using real-world data from one of the largest most ethnically diverse UK DESP we have shown marked sociodemographic differences in the development of STDR and PDR among PLD at low risk of diabetes related sight loss. Younger age groups (<45 years) compared with older age groups (especially those aged 55 to <65 years), and those of black and South Asian ethnic origin compared with whites, were at greater risk of developing STDR. Higher STDR among the youngest age group is of particular concern, given their trajectory for longer exposure to disease. Given these sociodemographic differences, we have shown that introducing biennial as opposed to annual diabetic eye screening could worsen sight loss among certain sociodemographic groups because of delayed detection of STDR and PDR, potentially adding to healthcare inequalities.

The UK NSC has recommended biennial screening among PLD with no DR on two consecutive screening visits, as the risk of progression to referable was considered low (∼0.7% per year) and cases would still be treatable if delayed.^8^ This recommendation was underpinned by a large study of 354,549 PLD from 7 nationally dispersed UK DESP,^9^ which showed confidence intervals of study estimates of referable diabetic eye disease ranging from 0 to 1.6%, and a calculated p-value for heterogeneity of <0.001. While heterogeneity was attributed to potential differences in age, sex, ethnicity and glycaemic control of screened populations, sociodemographic characteristics of the 7 studies were not outlined, and effects of sociodemographic factors were not explicitly quantified. While percentages of referable retinopathy were considered low, numbers will be considerable both in terms of delayed diagnosis and potentially irrecoverable sight loss when scaled up to the national screening programme.^3^ The current study explicitly quantified the potential impact of a biennial screening frequency by ethnic and age group, and identified those who would be more adversely affected. Moreover, we have previously shown that these high-risk sociodemographic groups, especially younger age groups, are less likely to attend screening appointments among this screened population.^15^ The introduction of biennial screening could plausibly disenfranchise PLD with no retinopathy from the programme, especially among the more disadvantaged or high risk sociodemographic groups, leading to further delays in diagnosis. A key issue is the potential adverse consequences of delayed diagnosis as a result of biennial screening. While those with referable retinopathy could still be treated later, as acknowledged within NSC recommendations,^8^ inevitably there would be more extreme cases (i.e., with PDR) who would experience irrecoverable sight loss. While we have shown that PDR occurrence is small, ethnic and age group disparities in the numbers are apparent and would still be appreciable within such a large screening programme, particularly among the oldest and black ethnic groups.

With increasing numbers being seen by the DESP ^3^ within a publicly funded healthcare system, the need for cost effectiveness while maintaining patient safety is paramount. The DESP could adopt the NSC recommendation for biennial screening among those at low risk, accepting that this would lead to age and ethnic inequality, or consider a more nuanced screening interval by sociodemographic factors to avoid inequality as suggested by others.^18^ However, tailored screening intervals would need to be decided and resources made available to administer such screening appointments. Alternatively, artificial intelligence (AI) technologies could be used to assist in maintaining the current status quo in screening frequency. Automated AI diabetic eye screening has been used in Scotland for over a decade, and is used or being considered for use elsewhere.^22^ However, AI screening is not currently licenced for use in the English NHS DESP, although a recent evidence synthesis review recommending staged implementation of one commercial system,^22^ which has been extensively validated to show adequate levels of screening performance and could halve the work load of human graders.^22-25^ Using AI to filter out images without DR has been shown to be safe and cost-effective,^24,25^ especially as humans could take longer to grade retinal images to ensure absence of retinopathy if a 2-year screening interval were to be adopted. While the effectiveness of AI has been demonstrated,^22-25^ quantifying equity of AI performance across different ethnic and age groups is needed; akin to formally assessing the potential impact of a biennial screening programme in different sociodemographic groups carried out in this study. Previous work has shown the potential cost-effectiveness of these different screening approaches, but further economic modelling is needed to directly compare cost effectiveness of these different approaches, particularly among less privileged sociodemographic groups.^5,6,18,26^

Our study has several strengths. First, use of a large, multi-ethnic DESP to determine incidence of STDR and PDR among PLD without retinopathy in different sociodemographic groups, particularly by ethnicity where there were high levels of recording (∼99%). While 41% of the cohort who had 2 consecutive screening episode without DR were used, prevalence of DR in the entire cohort was reassuringly similar to previous reports and is representative of the UK.^6,12,27^ DR classification was carried out by trained assessors within the NELDESP, following a multilevel internally and externally quality-assured grading protocol that meets national recommendations. Limitations include the use of annual screening data to simulate biennial screening. These findings may give an over optimistic indication of compliance as implementing biennial screening may worsen adherence to an extended screening regime. However, these findings using real-world data reflect clinical practice. A randomised controlled clinical trial would be the gold standard of assessing the impact of biennial screening, but such a study would need to be large to compare impact across different age and ethnic groups. More importantly, technologies to assist in screening are evolving so rapidly, findings could well be outdated before completion. Hence, we believe using ‘real-world’ large scale NHS data to assess the impact is important.

The incentive of biennial screening is to release capacity in the NHS and lessen the inconvenience for PLD at low risk of sight loss of attending eye screening appointments every year,^8^ but there is a need to address the potential to amplify ethnic and age inequalities in healthcare.^28^ This study is unique in providing the comprehensive high-quality data needed to inform policy-makers and healthcare professionals about potential age and ethnic ramifications of introducing a change in screening frequency, particularly in deprived populations.^29^ We would urge replication of these findings in other multi-ethnic DESP. Our findings suggest that ethnic and age inequalities in care could worsen with the introduction of biennial screening among PLD at low risk of diabetes-related sight loss. Moving forward either alternative technologies which could allow annual screening of PLD at low risk to continue, or more nuanced screening intervals among different sociodemographic groups warrant further consideration in providing more equitable healthcare.

## Data Availability

The North-East London Diabetic Eye Screening Programme data are not publicly available because of restrictions on data sharing. A fully anonymised data set is available from the Programme upon reasonable request.

## Competing interests

AO-B: none, ARR: none, CGO: none. JA: none. AW: none. YW: none. LB: none. RC: none. SB: none. JF: none. RW: none. PR: none. AH: none. RS: none. EYC: none. AYL: has received fees from Santen, Genetech, FDA, Johnson and Johnson, Carl Zeiss Meditec, Gyroscope, Regeneron, and has a non-remunerative relation with Microsoft. CE: has received fees from Heidelberg Engineering, Inozyme Pharma. AT: has received fees from Annexon, Apellis, Bayer, Genentech, Iveric Bio, Novartis, Oxurion, and Roche.

## Financial support

This work is funded by (i) Wellcome Collaborative Award (2022, 224390/Z/21/Z), and (ii) NHS Transformation Directorate and The Health Foundation, managed by the National Institute for Health and Social Care Research. The views expressed in this publication are those of the author(s) and not necessarily those of the NHS Transformation Directorate, The Health Foundation, National Institute for Health Research, or the Department of Health and Social Care.

## Author’s contributions

AO-B, CGO, ARR, JA, AT, and CE designed the study. JA, LB, and RC acquired the data. RC, JA, and AO-B curated the data. AO-B, CGO, and ARR undertook statistical analyses. AO-B, CGO, ARR, JA, and RC had direct access and verified the underlying data. AO-B and CGO wrote the first version of the manuscript. All authors, interpreted results, critically reviewed, and edited the manuscript. AO-B and CGO had the final responsibility for the decision to submit for publication. All authors had access to the data and have approved the decision to submit for publication.

## The Artificial Intelligence & Automated Retinal Image Analysis Systems (ARIAS) Research Group

John Anderson, Sarah Barman, Louis Bolter, Ryan Chambers, Lakshmi Chandrasekaran, Umar Chaudhry, Emily Y Chew, Catherine Egan, Jiri Fajtl, Frederick L Ferris, Aroon D Hingorani, Aaron Y Lee, Abraham Olvera-Barrios, Christopher G Owen, Paolo Remagnino, Alicja R Rudnicka, Royce Shakespeare, Reecha Sofat, Adnan Tufail, Alasdair N Warwick, Charlotte Wahlich, Roshan Welikala, Kathryn Willis, Yue Wu.

**Supplemental Figure 1:**
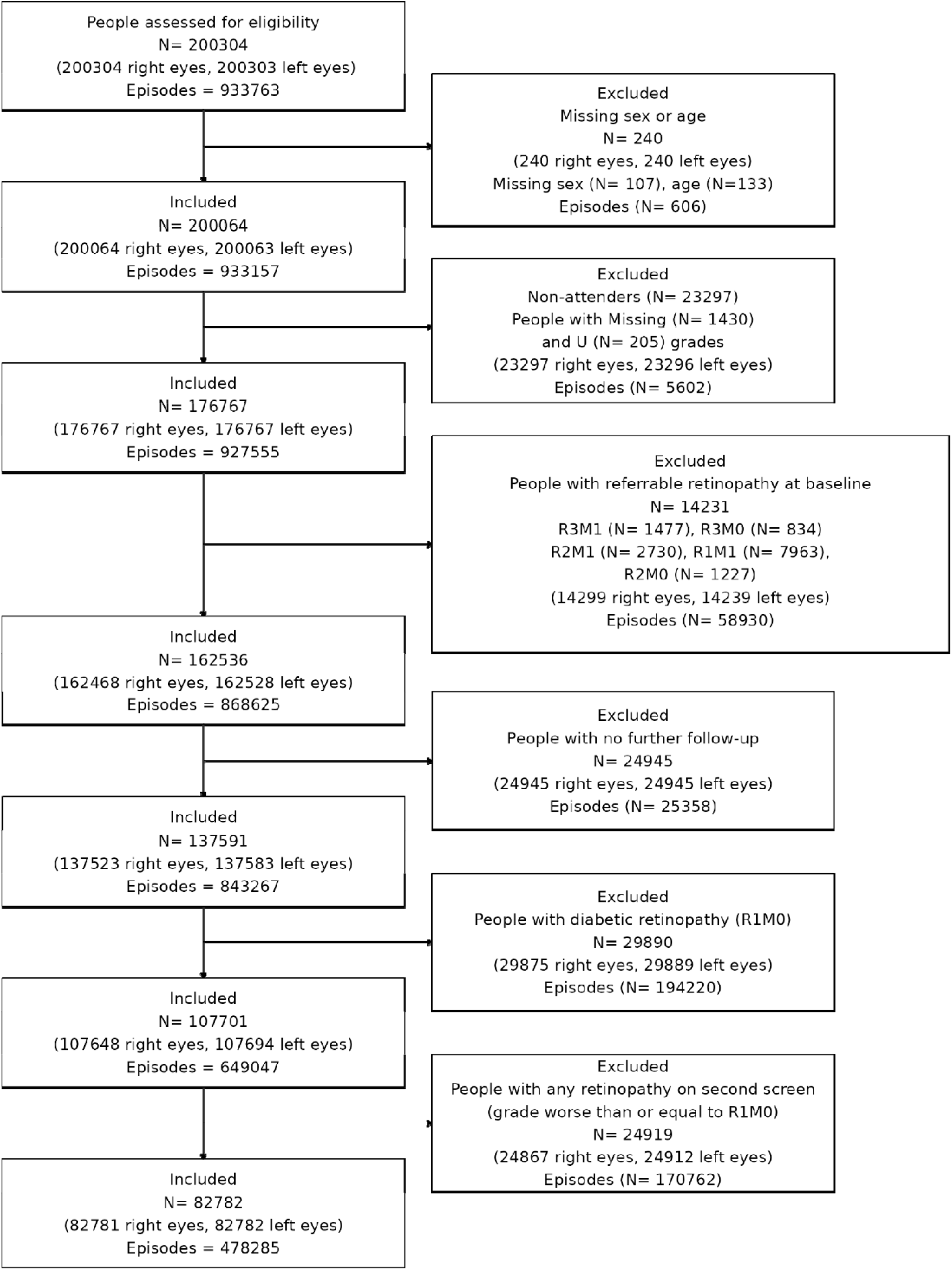
Diagram of exclusions.

**Supplemental Table 1:** Cumulative incidence rates (%) of STDR among PLD without diabetic retinopathy on 2 consecutive screening appointments, overall by age groups, sex, ethnic group and IMD N= 82,782.

| Characteristic | Year 1 | Year 2 | Year 3 | Year 4 | Year 5 | Year 9 |
| --- | --- | --- | --- | --- | --- | --- |
| **Overall** | 0.20 (0.17 - 0.22) | 0.33 (0.30 - 0.36) | 0.40 (0.36 - 0.43) | 0.44 (0.40 - 0.48) | 0.45 (0.41 - 0.49) | 0.51 (0.47 - 0.55) |
| **Age groups** |  |  |  |  |  |  |
| Less than 45 years | 0.19 (0.13 - 0.25) | 0.35 (0.28 - 0.43) | 0.44 (0.35 - 0.52) | 0.48 (0.39 - 0.57) | 0.52 (0.42 - 0.61) | 0.67 (0.56 - 0.77) |
| 45 to <55 years | 0.18 (0.13 - 0.23) | 0.31 (0.25 - 0.38) | 0.36 (0.29 - 0.43) | 0.41 (0.33 - 0.48) | 0.42 (0.35 - 0.50) | 0.49 (0.41 - 0.57) |
| 55 to <65 years | 0.17 (0.13 - 0.22) | 0.27 (0.21 - 0.33) | 0.32 (0.26 - 0.38) | 0.35 (0.28 - 0.42) | 0.35 (0.29 - 0.42) | 0.39 (0.32 - 0.46) |
| 65 years and over | 0.24 (0.19 - 0.29) | 0.38 (0.31 - 0.44) | 0.47 (0.40 - 0.54) | 0.51 (0.44 - 0.58) | 0.51 (0.44 - 0.59) | 0.53 (0.45 - 0.60) |
| **Sex** |  |  |  |  |  |  |
| Female | 0.22 (0.19 - 0.26) | 0.35 (0.30 - 0.40) | 0.44 (0.38 - 0.49) | 0.48 (0.42 - 0.54) | 0.48 (0.42 - 0.53) | 0.54 (0.48 - 0.60) |
| Male | 0.17 (0.14 - 0.21) | 0.31 (0.27 - 0.35) | 0.36 (0.31 - 0.41) | 0.40 (0.35 - 0.45) | 0.42 (0.37 - 0.47) | 0.48 (0.43 - 0.54) |
| **Ethnicity** |  |  |  |  |  |  |
| White | 0.15 (0.12 - 0.19) | 0.22 (0.18 - 0.27) | 0.27 (0.22 - 0.32) | 0.29 (0.24 - 0.35) | 0.30 (0.25 - 0.35) | 0.34 (0.29 - 0.40) |
| South Asian | 0.21 (0.17 - 0.26) | 0.37 (0.31 - 0.42) | 0.43 (0.37 - 0.49) | 0.47 (0.41 - 0.54) | 0.48 (0.42 - 0.55) | 0.55 (0.48 - 0.62) |
| Black | 0.26 (0.18 - 0.33) | 0.47 (0.37 - 0.57) | 0.59 (0.49 - 0.70) | 0.67 (0.56 - 0.79) | 0.71 (0.59 - 0.83) | 0.77 (0.65 - 0.90) |
| Any other Asian | 0.23 (0.12 - 0.35) | 0.29 (0.16 - 0.42) | 0.39 (0.24 - 0.53) | 0.44 (0.28 - 0.60) | 0.43 (0.28 - 0.59) | 0.48 (0.31 - 0.64) |
| Other | 0.26 (0.09 - 0.43) | 0.40 (0.18 - 0.61) | 0.41 (0.19 - 0.63) | 0.47 (0.23 - 0.70) | 0.48 (0.25 - 0.72) | 0.63 (0.36 - 0.90) |
| Mixed | 0.20 (0.00 - 0.43) | 0.60 (0.20 - 1.00) | 0.69 (0.26 - 1.12) | 0.61 (0.21 - 1.02) | 0.54 (0.16 - 0.93) | 0.67 (0.25 - 1.10) |
| Chinese | 0.00 (0.00 - 0.00) | 0.10 (0.00 - 0.31) | 0.21 (0.00 - 0.52) | 0.23 (0.00 - 0.56) | 0.25 (0.00 - 0.60) | 0.21 (0.00 - 0.52) |
| Unknown | 0.16 (0.00 - 0.42) | 0.67 (0.14 - 1.19) | 0.59 (0.09 - 1.08) | 0.55 (0.07 - 1.02) | 0.52 (0.06 - 0.99) | 0.50 (0.05 - 0.96) |
| **Type of diabetes** |  |  |  |  |  |  |
| Type 2 | 0.20 (0.17 - 0.22) | 0.32 (0.29 - 0.35) | 0.39 (0.35 - 0.43) | 0.43 (0.39 - 0.47) | 0.44 (0.40 - 0.48) | 0.50 (0.46 - 0.54) |
| Type 1 | 0.38 (0.16 - 0.60) | 0.53 (0.27 - 0.79) | 0.67 (0.38 - 0.96) | 0.75 (0.44 - 1.05) | 0.77 (0.46 - 1.09) | 0.84 (0.52 - 1.17) |
| Other | 0.00 (0.00 - 0.00) | 0.42 (0.00 - 1.33) | 0.96 (0.00 - 2.34) | 0.82 (0.00 - 2.09) | 0.75 (0.00 - 1.96) | 0.86 (0.00 - 2.15) |
| **Index of Multiple Deprivation** |  |  |  |  |  |  |
| 1 | 0.16 (0.09 - 0.23) | 0.25 (0.16 - 0.33) | 0.36 (0.26 - 0.47) | 0.43 (0.31 - 0.54) | 0.45 (0.34 - 0.57) | 0.48 (0.36 - 0.61) |
| 2 | 0.22 (0.17 - 0.27) | 0.37 (0.30 - 0.43) | 0.43 (0.36 - 0.49) | 0.47 (0.40 - 0.54) | 0.48 (0.41 - 0.55) | 0.54 (0.47 - 0.62) |
| 3 | 0.17 (0.12 - 0.21) | 0.32 (0.26 - 0.38) | 0.40 (0.34 - 0.47) | 0.43 (0.36 - 0.50) | 0.43 (0.36 - 0.50) | 0.50 (0.43 - 0.58) |
| 4 | 0.25 (0.18 - 0.31) | 0.34 (0.27 - 0.42) | 0.38 (0.30 - 0.46) | 0.42 (0.34 - 0.51) | 0.43 (0.35 - 0.52) | 0.49 (0.40 - 0.58) |
| 5 | 0.17 (0.10 - 0.25) | 0.31 (0.21 - 0.41) | 0.36 (0.25 - 0.47) | 0.41 (0.29 - 0.53) | 0.42 (0.30 - 0.54) | 0.48 (0.35 - 0.60) |

## Notes

### Author Declarations

The study was approved by the NHS Health Research Authority and Health and Care Research Wales (IRAS No. 265637). The study was carried out in accordance with the Declaration of Helsinki.

